# Hydroxyurea Therapy for Neurological and Cognitive Protection in Pediatric Sickle Cell Anemia in Uganda (BRAIN SAFE II): Protocol for a single-arm open label trial

**DOI:** 10.1101/2024.01.12.24301208

**Authors:** Vincent Mboizi, Catherine Nabaggala, Deogratias Munube, John M. Ssenkusu, Phillip Kasirye, Samson Kamya, Michael G. Kawooya, Amelia Boehme, Frank Minja, Ezekiel Mupere, Robert Opoka, Caterina Rosano, Nancy S. Green, Richard Idro

## Abstract

**Background:** Children with sickle cell anemia (SCA) in Sub-Saharan Africa are at high risk of sickle cerebrovascular injury (SCVI). Hydroxyurea, a commonly used disease-modifying therapy, may prevent or decrease SCVI for reduced incident stroke, stroke risk and potentially cognitive dysfunction. We aim to test the impact of daily hydroxyurea therapy on these outcomes in Ugandan children with SCA. We hypothesize that hydroxyurea therapy over 36 months will prevent, stabilize or improve these complications of SCA.

**Methods:** The BRAIN SAFE II study is an open-label, single-arm trial of daily hydroxyurea for 270 children with SCA (HbSS) in Uganda, ages 3-9 years. Following baseline assessments, participants began hydroxyurea therapy and clinically followed per local guidelines. Standard hydroxyurea dose is escalated to maximum tolerated dose (MTD). SCVI is assessed by cerebral arterial velocity using Doppler ultrasound, with cognitive function determined by formal neurocognitive testing (primary outcomes). Structural SCVI is assessed by magnetic resonance imaging (MRI) and angiography (MRA) in a sub-sample of 90 participants ages ≥5 years, along with biomarkers of anemia, inflammation and malnutrition (secondary outcomes). At trial midpoint (18 months) and completion (36 months), primary outcomes will be compared to participants’ baseline to determine hydroxyurea impact and relationships to secondary outcomes.

**Conclusion:** This open-label, single-arm trial will examine the impact of hydroxyurea on preventing or ameliorating SCA SCVI in children, assessed by reducing incident stroke, stroke risk and neurocognitive dysfunction. Trial results will provide important insight into the role of hydroxyurea therapy on critical manifestations of SCVI in children with SCA.

## BACKGROUND

Annually, over 400,000 babies are born with Sickle Cell Anemia (SCA) worldwide, with most born in sub-Saharan Africa.(1) Approximately 20,000 infants, 1-2% of births, are born with SCA annually in Uganda.(2) Disease is characterized by chronic illness and repeated acute symptoms, e.g. severe pain. Affected persons are at high risk of stroke due to development of sickle cerebrovascular injury (SCVI).(3-5) SCVI also can lead to neurocognitive impairment from accumulated sub-clinical infarcts.

Extent of stroke risk is stratified by cerebral arterial flow velocity measured by transcutaneous doppler ultrasound (TCD). In high-resource regions, primary stroke reduction has been achieved through TCD screening to institute stroke prevention therapy for those at high risk.(6) TCD screening is not available to most people with SCA in Africa.(7) Our meta-analysis estimated that as many as 60,000 children in sub-Saharan Africa have SCA stroke, emphasizing the high burden in vulnerable children.(8) In addition, clinically unapparent stroke can only be detected by magnetic resonance imaging and arteriography (MRI-MRA).(5, 9)

Our prior cross-sectional study of neurological and neurocognitive abnormalities of children with SCA in Uganda, “Burden of Neurological and Neurocognitive Impairment in Pediatric Sickle Cell Anemia in Uganda (BRAIN SAFE)” found that over 20% had ≥1 impairment.(10) Based on those findings, we plan to conduct a trial to test the impact of hydroxyurea therapy in children with SCA on the frequency and severity of SCVI manifestations.

Hydroxyurea reduces stroke risk in children with SCA, largely through increased hemoglobin and fetal hemoglobin levels, leading to improved blood flow for cerebral oxygen availability.(5) These physiological changes are detected by improved TCD arterial blood flow velocities.(5, 11, 12) However, the full effects of hydroxyurea on SCVI in children have not yet been established in low- and middle-income countries (LMICs) in the sub-Saharan region (Figure 1). In high-income countries, chronic blood transfusions is the initial treatment for primary stroke prevention in children with abnormal TCD, with later transition, if meeting established criteria, to hydroxyurea therapy.(13) However, safe, reliable chronic transfusions are not feasible in many lower-resource settings, including sub-Saharan Africa. More recently, some sub-Saharan studies of hydroxyurea have demonstrated improved TCD measurements and reduced stroke risk.(12, 14)

BRAIN SAFE II is a single arm, open label 36-month trial of daily oral hydroxyurea therapy on the frequency and severity of SCVI, compared to baseline, in a cohort of children treated at the Mulago Hospital SCA clinic (MHSCC) in Kampala, Uganda. Our hypothesis is that hydroxyurea therapy will improve or stabilize SCVI, as measured by incident stroke, TCD velocity, neurocognitive testing, and – for a 30% sub-sample – stabilize MRI-MRA findings of cerebral infarcts and arterial stenoses.

The primary objective is to test the hypothesis, assessing stability or improvement compared to baseline in each of these 3 age-adjusted outcomes, as well as the age-adjusted findings of our prior cross-sectional study performed at MHSCC.(10) Secondary objectives are to evaluate the impact of hydroxyurea therapy on structural SCVI using MRI and MRA in a cohort subset and to determine if measures of known or suspected risk factors of SCVI, specifically anemia, inflammation, as measured by blood levels of C-reactive protein (CRP), and malnutrition, are associated with stabilized or improved primary outcomes, and with the secondary outcome detected by MRI-MRA.

## METHODS

### Ethics approval and consent

This protocol and all other participant-facing study documents were approved by the Makerere University School of Medicine Research and Ethics Committee (SOMREC protocol #2019-147) and Columbia University (Columbia protocol AAAS8955). Administrative clearance was received from Mulago National Referral Hospital (MHREC# 1802), Uganda National Council of Science and Technology (HS864ES) and the National Drug Authority (CTA0147).

### Study location

Mulago Hospital sickle cell clinic (MHSCC), a well-established specialized clinic at the Mulago National Referral Hospital, is staffed by pediatricians and pediatric hematologists in collaboration with the Makerere University College of Medicine.(10, 15) MHSCC is the largest SCA clinic in Uganda, providing cost-free specialized care to affected children. Over 5000 pediatric patients are actively followed, with 350-450 weekly visits.(16) The clinic offers standard of care for SCA in accordance with the Uganda Ministry of Health national SCA guidelines, including monthly supplies of folic acid, malaria prophylaxis and, for children to the age of age 5 years, penicillin V.(17)

### Eligibility criteria and informed consent

Based on MHSCC records, parents/guardians of children who met the screening eligibility criteria (Table 1) were contacted and invited to the study clinic to provide written consent. Among the criteria, enrolment required the absence of evidence of prior stroke detected by a standardized stroke examination, PedNIHSS.(10) Consent (and assent for children ages ≥8 years) for trial procedures were provided in English or a local language (Luganda), per parent/guardian and participant preference.

### Intervention

This is an open-label single arm trial. Hydroxyurea is a well-studied medication for children with SCA for reducing disease-associated complications.(18-20). Hydroxyurea therapy increases hemoglobin levels and induces a dose-dependent increase in fetal hemoglobin (HbF) levels. HbF, a minor component of total hemoglobin, reduces hemolysis and anemia by inhibiting the polymerization of sickle hemoglobin, and thereby the sickling of red blood cells. Other hematological effects include reduced numbers of white blood cells and platelets, thought to be useful for decreasing vascular activation and inflammation.

An earlier Ugandan pediatric trial demonstrated hydroxyurea safety in children with SCA, despite endemic malaria, as well as its efficacy in reducing acute pain crises and need for blood transfusions.(21) This study led to the Uganda approval of hydroxyurea for children. More recently, a Ugandan randomized trial of standard hydroxyurea dose versus dose escalation demonstrated clinical advantage to the latter.(22) Due to these established benefits, all the participants in the BRAIN SAFE II trial were started on hydroxyurea with planned dose escalation.

### Study drug and dosing

Study drug is a hydroxyurea product, Siklos® (AddMedica/Theravia, France). It is formulated for accurate pediatric dosing and swallowing in readily dispersible scored tablets of 100 and 1000mg. A gradual escalation to maximum tolerated dose (MTD) up to 30mg/kg/day is planned, based on current standard international guidelines for escalation and haematological toxicity, especially myelosuppression (Table 1).(22) Dose escalation is planned for all participants, regardless of laboratory or clinical improvement on lower doses.

- Hydroxyurea dose escalation will occur in 2.5 mg/kg/day increments, adjusted every 8 weeks unless a haematological toxicity occurs or reaches MTD;
- Target absolute neutrophil count (ANC) on hydroxyurea therapy is 2.0 - 4.0 x 10^9^/L, similar to goals of a previous hydroxyurea clinical trial.(22) Dose-limiting MTD may also be based on low absolute reticulocyte count (ARC).

### Strategies for drug adherence

Residual pill counts are performed by study staff at each study visit. Adherence is also assessed by parental/guardian report of adherence and by haematological laboratory evidence of reduced dose exposure, e.g. rising neutrophil counts and reduced mean corpuscular volume (MCV) compared to earlier changes.

Participants with excess residual study tablets and the above evidence are identified as less adherent than study requirements. Staff will provide counselling for adherence to parents/guardians and work to identify addressable adherence barriers. Caregivers will be reminded not to share study drug with other family members with SCA.

### Criteria for discontinuing trial participation

Participants may be removed from the trial by the Principal Investigators (PIs) for any of these reasons: severe allergic reaction or life-threatening drug-related toxicity; withdrawal of approval by the parent/guardian; participants who miss two scheduled study visit in any 12-month period; participant withdrawal for poor adherence despite staff interventions.

### Medical care during the trial

In addition to the study drug, parents/caregivers are educated on the care of children with SCA, including hydration, care during cold weather, pain management and when to seek medical help. Hospitalizations for SCA complications and other serious medical events will use MHSCC admission criteria.

### Sample size

Sample size of 270 participants will provide sufficient power (alpha=0.05; beta=84%) to detect decreased arterial TCD velocity by 15 cm/second over 3 years in children with abnormally elevated velocities at baseline.(23) This difference is clinically significant, and sufficient to move a child into or towards the normal range, thereby lowering stroke risk.(24) Sample size determinations included an additional 8% to accommodate expected losses due to stroke, death or other loss to follow-up. The proposed sample size is also expected to provide sufficient power to detect even modest changes in cognition over time.

### Recruitment

We randomly selected, consented, and enrolled a cohort of 270 children ages 3-9 years from among eligible patients attending MHSCC. Children ≥8 years were asked for assent. Final screening included a neurological exam using the PedNIHSS (Figure 2).(25, 26) Eligibility required lack of with evidence of prior stroke by a standardized pediatric stroke scale (PedNIHSS). Participants are provided with and monitored on hydroxyurea for the entire study period. Most scheduled study assessments will be those we have previously described.(10)

A sample of 110 siblings or close family relatives without SCA were recruited as controls and to create age-specific z-scores for neurocognitive testing, malnutrition and other study assessments. Controls were recruited at each expected participant age group throughout the trial: ages 3-12 years. Eligibility required lack of neurologic evidence of prior stroke PedNIHSS or other neurologic condition; and confirmed lack of SCA by hemoglobin electrophoresis.

### Implementation

Baseline assessments consisted of a detailed history, including age at SCA diagnosis, number and reasons for previous hospitalizations, blood transfusions and severe SCA complications. In addition, standard anthropometric, neurologic, TCD and neurcognitive testing, and neuroimaging testing, complete blood count and renal and hepatic function tests were obtained (Figure 2, Tables 2, 3). Subsequently, scheduled visits include a general health assessment, weight and blood count monitoring for study drug safety and ongoing weight-based dosing.

After completing baseline assessments, participants were initiated on daily hydroxyurea therapy of 20+/-2.5mg/kg/day. Families of enrolled children are provided with dose instructions and sufficient study drug supply until the next scheduled visit. Dose escalation is performed at scheduled visits and according to standardized safety consideration for blood counts (described below).(22). In addition, periodic dose adjustments are performed for weight changes to maintain stable daily per-kilogram intake. Potential hematological, hepatic and renal toxicities are monitored prior to dosing and at annual intervals (Table 3).

### Primary outcomes

The primary efficacy outcome measures are the frequency, age-specific prevalence and severity of SCVI manifested by >1 of the 3 end points after 3 years compared to baseline assessments (Figure 1, Table 4) and the age-specific prevalence of these outcomes determined by our prior cross-sectional study, BRAIN SAFE (10):

a. Reduced maximal TCD velocity by >15cm/second and by improved TCD using standard categorical assessment - normal, conditional or abnormal - for children with SCA; (27)
b. Stabilized or improved cognitive impairment using age-specific z-scores and categorical (impaired/not impaired) using standard criteria of z-scores ≤-2 using compared to baseline;
c. New stroke, as assessed by PedNIHSS, or death where stroke appears to be the inciting event. Rate of incident stroke will be compared to reports of children with SCA in the region, including the 5-month period of our prior cross-sectional study at MHSCC. (10)

### Secondary outcomes

Secondary outcome measures are:

a. Proportion of the sub-sample with magnetic resonance imaging who have abnormalities detected (yes/no); and stabilized or worsened structural changes on brain magnetic resonance and angiography imaging (MRI and MRA) at the follow-up scan at 36 months compared to baseline.(10, 28)
b. Higher haemoglobin and HbF, lower CRP levels
c. And, improved nutrition status by WHO standards (the latter by age-adjusted international z-score) and by bioimpedance analysis (BIA) to assess changes in body composition at months 18 and 36 compared to baseline values.(10, 29)

### Measurement of outcomes

Assessment for all 3 primary endpoints are scheduled for enrolment, months 18 and 36 (Figure 2) and (supplementary Table 1):

#### Incident stroke

Frequency of new strokes will be assessed as the event per 1000 patient years. Where possible by hospitalization at Mulago, children with an acute neurological event are evaluated by the PedNIHSS for stroke confirmation.

#### TCD velocity

The arterial time-averaged maximum mean velocities (TAMV) for each of seven ajor cerebral vessels will be acquired.(30) High stroke risk with elevated flow velocities, defined by the standardized criteria for pediatric SCA as conditional (≥170-<200 cm/second) or abnormal (≥200 cm/second) in one or more arteries tested, predicts high SCA stroke risk in children ages 2-16 years.(30, 31) Participants with abnormal TCD at enrolment and/or month 18 will in addition be offered blood transfusions to reach a haemoglobin level of 9-10 grams/dL. TCD will be repeated TCD after 3 months to re-assess TAMV, and every 6 months until stable.

#### Neurocognitive function

SCA participants undergo standardized age-appropriate test batteries. All test materials were previously translated into and validated in Uganda, with participant choice of test language.(10) Neurocognitive domains tested are cognitive ability, attention and executive function, as all three can be impaired in children with SCA.(32) Children with z-scores below -2 SD from reference norms are considered as impaired. Practice effects from multiple testing is minimized by the gap in time between baseline testing and study schedule.(33, 34)

Cognition is assessed for all participants using the Kaufman Assessment Battery for Children, 2nd edition (KABC-II). The KABC-II has five subscales: sequential processing, simultaneous processing, learning, planning and knowledge.(35, 36). Attention will again be measured using the laptop-based Test of Variables of Attention (TOVA).(10, 37)

For executive function, children ages 3 to <5 years will be assessed using the Behavioral Rating Inventory of Executive Functions, Preschool edition (BRIEF-P), a questionnaire completed by the primary caregiver.(38) Children ages ≥5 are tested for executive function by the school-age version of BRIEF.(39) In addition, executive function for those >6 is also assessed by the Developmental Neuropsychological Assessment (NEPSY), second edition (NEPSY-2).(40) This testing platform assesses various cognitive domains in children 3 to 16 years, has good psychometric properties and has been used in South African studies.(41, 42)

### Secondary outcomes

#### Laboratory evidence of improved SCA status

The main impact of hydroxyurea on SCA is through its improved anemia, higher HbF levels and reduced inflammation. Secondary outcomes include degree of elevation of haemoglobin and HbF levels, and reduced CRP.

#### Growth and nutritional status

are assessed using height- and weight- for age (ages <5 years) or BMI for age z scores (≥5 years) using WHO standards for malnutrition for sex and age.(10, 43) Malnutrition is defined by with -2 z-scores or lower.(10) Additional evaluation of the impact of hydroxyurea on malnutrition is assessing using a non-invasive assay of body composition for estimating longitudinal changes. We are using multi-frequency bioelectrical impedance analysis (BIA) (QuadScan MODEL Vacumed, Ventura, CA USA), which has previously been used for estimating body composition of children in the region.(44) Specifically, BIA provides an estimate of lean body weight (fat mass (FM) and fat-free mass (FFM) using our controls’-generated gender- and age-specific standards.

#### MRI-MRA

Scans are performed on a 30% subset of participants ages ≥5 at baseline and study completion. Standard non-contrast imaging is performed using a 1.5 tesla scanner (1.5T Achieva MRI equipment, Philips Medical Systems, Netherlands)(10). The age limit aimed to include children able to be scanned without motion artifact despite lack of sedation. Standard clinical interpretation is performed by one of two study radiologists who will be blinded to other study results.(10) A blinded research interpretation is performed by an independent U.S. neuro-radiologist.(10) All disagreements between the two reads will be adjudicated by a third, blinded independent neuro-radiologist.

### Data collection and management

Participants were assigned a non-identifying numeric code for secure de-identification to maintain confidentiality for all study records. Data are collected and retained using both paper-based and electronic record forms. The password-protected study database uses REDCap (https://www.project-redcap.org), hosted by Global Health Uganda with cloud back-up. Paper records are kept in a locked file cabinet in the Uganda PI’s office. Data will be kept electronically in compliance with prevailing laws on data storage.

### Participant retention

Participants are reimbursed for round-trip transportation costs to clinic for enhanced visit attendance. Contact information/telephone and location and directions to their homes are recorded. Caregivers are called if children miss scheduled follow-up visit.

### Laboratory evaluations and storage of biological specimens

Table 3 summarizes the schedule and specifics of laboratory testing at enrolment and scheduled assessments.

a. At enrolment, laboratory testing included a full blood count, HbF level and safety labs. At subsequent study visits, safety labs are performed in accordance with the MHSCC clinical protocol.
b. Additional plasma samples are obtained at time 0, 18 months and 36 months for CRP testing and storage for biomarker testing.
c. Cell pellets from the plasma sample will be stored for future DNA testing for SCA variants and other polymorphisms.

### Patient characteristics and baseline comparisons

At enrolment, descriptive statistics will characterize baseline demographic and clinical features. These characteristics shall include the child’s age, sex, anthropometric measurements, mother’s (or primary caregiver’s) age and educational attainment and family socioeconomic status. Caregiver questionnaire reports SCA complications, serious infections and hospital admissions in the pretrial 12 months, haemoglobin level. For each primary outcome, baseline characteristics of children who did/did not experience an event an experience an event will be compared using a Student’s t- and Chi-squared tests. Non-parametric tests shall be used as needed. Variables that show an association with the occurrence (or non-occurrence) of the events (P<0.2) will be selected for the multivariable analysis in the multivariable models for the respective outcomes.

### Primary analysis

The primary outcome analysis will include neurological examination for incident stroke, TCD and neurocognitive outcomes as distinct endpoints to gauge the effect of hydroxyurea at subsequent time points (18 and 36 months) compared to baseline measures performed by intention to treat. In addition, analysis of participants who reach an early endpoint of stroke or death, thereby terminated the trial early, will be compared to the remaining sample. Crude and adjusted mixed effects models will be adjusted for baseline characteristics with P<0.2 at bivariate analyses. Generalized and linear mixed effects models (GLMM) account for the correlation of repeated measures on the same child. Unadjusted analyses shall be conducted, followed by adjusted analyses shall be conducted by adding other baseline characteristics with P<0.2 to the models in addition to the time variable. We will treat age as a time-varying covariate to account for age differences. No data imputation will be conducted for missing data.

### Assessment of efficacy

TCD shall be measured both as a quantitative (by arterial flow velocity) and as categorical (normal vs. conditional/abnormal) variable. We will use linear mixed effects models (LMM) to assess within and between participant changes over time from month 0 to 36 for the continuous TCD velocity measure, where participant-level variation will be captured by a participant-specific random effect.

Neurocognitive tests shall also be both quantitative (z-scores) and categorical (less than -2 z-scores vs. -2 z-scores and above, defining neurocognitive deficits present or absent, respectively. Changes in binary neurocognitive measures over time will be modelled using GLMM.

Incident stroke will be determined through NIHPedSS, to be dichotomized into yes/no at each measurement time point (baseline 0, 18, and 36 months), considering time to the stroke. The proportional hazards assumption shall be evaluated both using log-log plots and Schoenfeld residuals and by testing the significance of interaction terms of model variables with time. The proportional cause-specific hazard assumptions shall also be assessed using Schoenfeld residuals.

### Analysis of secondary outcomes

Levels of haemoglobin, HbF, CRP and malnutrition status (the latter by z-score) over time will be compared to baseline measurements.

Brain magnetic resonance and angiography imaging (MRI-MRA) will be performed on a subset of 90 SCA participants (33%) at baseline and study month 36. We will assess imaging markers for SCVI at enrolment and subsequently for evidence of stabilization or worsening treatment over time(28). Changes in cerebral infarcts and vascular stenoses will be assessed as the proportion of: a) additional participants with infarct or stenoses; b) additional or larger/more severe infarcts or stenoses at trial completion for those with these abnormalities are baseline. Outcomes will be tested as a dichotomous outcome of MRI-MRA abnormality yes/no and stable or worse, using standardized SCA cerebral vasculopathy evaluations to identify patients whose scans worsen. Logistic regression will be used to identify risk factors such demographic, clinical and laboratory results for worsening versus stable imaging results.

### Changes in anaemia, CRP, and malnutrition status

Changes will be assessed for haemoglobin, CRP, and malnutrition z-scores at months 0, 18 and 36. To estimate hydroxyurea effects for each endpoint, we will use linear mixed effects models to assess within and between participant changes over time for the continuous measures of sickle cerebrovascular injury biomarkers (e.g. haemoglobin, CRP, and malnutrition z-scores), where participant-level variation is captured by random effects.

### Oversight and monitoring

All adverse events (AEs) and serious adverse events (SAEs) will be recorded, tabulated by type, intensity, seriousness, duration, and relationship to study drug and study procedure. Oral hydroxyurea is currently standard of care in Uganda for abnormal TCD or secondary stroke prevention in children with SCA. Moreover, its safety for this Ugandan population was demonstrated.(45) Hydroxyurea is used widely as standard of care in the U.S., France and elsewhere for children with SCA.(18, 46) For these reasons, interim safety analysis and futility testing were not required. Nonetheless, we constituted a Data Safety and Monitoring Committee (DSMC) to review any safety issues that may emerge compared to local standard of care. Its trial functions are to assure correct study procedures and participant safety. The independent members and the study biostatistician are each connected to SCA and clinical trials. The DSMC meets regularly at least twice annually to evaluate study progress and safety. All adverse events (IAEs) are documented following GCP principles. All severe AEs (SAEs) are promptly reported to the sponsor, DSMB and ethics committees within seven days of the investigators becoming aware of the event.

All scientific decisions are made jointly by the two PIs. Global Health Uganda oversees compliance with “Good Clinical Practice” (GCP), trial scientific integrity and oversight for all procedures, including data collection. Regularly scheduled communication, as well as *ad hoc* communication among the study team leadership supports high-quality trial conduct and quality assurance. Internal institutional compliance monitoring is conducted by Makerere University School of Public Health Clinical Trials Unit. External monitoring is conducted by the East African Consortium for Clinical Research Monitors.

## CONCLUSION

The high burden of SCVI, compounded by health system challenges in LMICs like Uganda, expose these children to excess morbidity, disability and mortality.(19, 47, 48) Here we evaluate the best available therapy for prevention and treatment of pediatric SCVI and continue to build capacity for leading collaborative multi-disciplinary research. As population-based newborn screening for SCA expands in Uganda and elsewhere in the region, this trial may facilitate the introduction of early SCVI prevention efforts.(49)

In addition to reduction of disease complications, e.g. pain and anemia, hydroxyurea clearly lowers TCD velocities among children with SCA.(4, 50) However, the extent to which hydroxyurea would improve or stabilize the cognitive function and structural abnormalities detected by MRI-MRA in sub-Saharan children with SCA remains unknown. The 3-year BRAIN SAFE II trial seeks to evaluate the effect of optimally dosed hydroxyurea on SCVI, manifested in both neurological and neurocognitive function.

### Trial status

Trial enrollment occurred from April to December 2021, following a 1-year delay due to COVID-19 pandemic starting in March 2020. Staff followed study-specific standard operating procedures (SOPs) to minimize risk of viral infection for staff, participants and their families. Baseline clinical assessments and initiation of hydroxyurea treatment for all participants commenced at or shortly after enrollment. An unselected subset of 90 ages ≥5 underwent MRI-MRA imaging.

Participants continue to undergo per protocol monitoring and study procedures. As of July 2023, all trial participants had completed study month 18, when a optional midpoint analysis was performed. Based on the month-18 interim analyses, Makerere Institutional Review Board (IRB) and the DSMC approved a shortening of the trial from 36 to 30 months. The 12-month following the midpoint assessment would permit assessment of continued or even further benefit from study drug. Further, a 12-month gap between neurocognitive testing for children is considered the minimum time to avoid practice effects from repeated testing.

## Data Availability

This is a methods paper. However, all data produced in the present work are contained in the manuscript

## Abbreviations

AE: Adverse event
AIDS: Acquired immunodeficiency syndrome
BRAIN SAFE: Hydroxyurea for the prevention of neurologic and cognitive impairment in children with sickle cell anaemia
CRF: Case Report form
CRO: Contract Research Organisation
DSMC: Data Safety and Monitoring Committee
GCP: Good Clinical Practice
HIV: Human Immuno-deficiency virus
IEC/IRB/REC: Independent Ethics Committee / Institutional Review Board
ITT: Intention to treat
ICH: The International Conference on Harmonisation
SD: standard deviation
NfLC: Neurofilament light chain
SOP: standard operating procedure
SP: Sulfadoxine-pyrimethamine
SSA: Sub-Saharan Africa
URTI: Upper respiratory tract infection
RBC: Red Blood Cells
SAE: Serious Adverse Event
WHO: World Health Organization

## Acknowledgements

We acknowledge AddMedica/Theravia for providing the intervention drugs as a donation. We also thank Global Health Uganda (GHU) for their input and trial sponsorship.

Study sponsor: Global Health Uganda (GHU).

P.O. Box 33842, Kampala, Uganda.

Tel: +256393516707

## Authors’ contributions

Author roles were: conceptualization (VM, RO, NSG, RI); Data curation (VM, CN, JMS); Formal analysis (JMS, AB); Funding acquisition (NSG, RI); Investigation (VM, DM, PK, SK, MGK, FM); Methodology (VM, CN, DM, EM, CR, NSG, RI); Project administration (VM, DM, PK, NSG, RI); Roles/Writing - original draft (VM, CN, NSG, RI); and Writing - review & editing (VM, NSG, RI).

## Funding

Funding to conduct the trial is provided by a grant from the Eunice Kennedy Shriver National Institute of Child Health and Human Development (NICHD) and the Fogarty International Centre at the National Institutes of Health (NIH) (1R01HD096559, Idro, Green).

Neither the sponsor nor funders had any role in the design of this trial and will not have any during the execution, analysis, interpretation of the data, or decision to submit the results. Per standard NICHD requirements for clinical trials as of 2019, the funder’s role is limited to periodic monitoring of the project timeline, developed by the study Principal Investigators, to help ensure timely completion of study procedures.

## Availability of data and materials

The full study protocol, supporting documents and the fully anonymised research database will be made publicly available once the study findings have been published. The data manager and statistical programmer will produce a document summarising the methods used to generate the data with a full description of all procedures, analyses, data capture tools, coding and description of variables. This document will be available alongside the research database.

In addition, per Release and Sharing of Data, our trial data sharing plan will follow the guidance for release to particular parties with restrictions rather than release for public use without restrictions. The agreement will include attention to issues related to co-authorship, reviews of findings produced through using the data, reports published about those findings, and the date the data are to be returned or destroyed.

---

**Table A.1.**
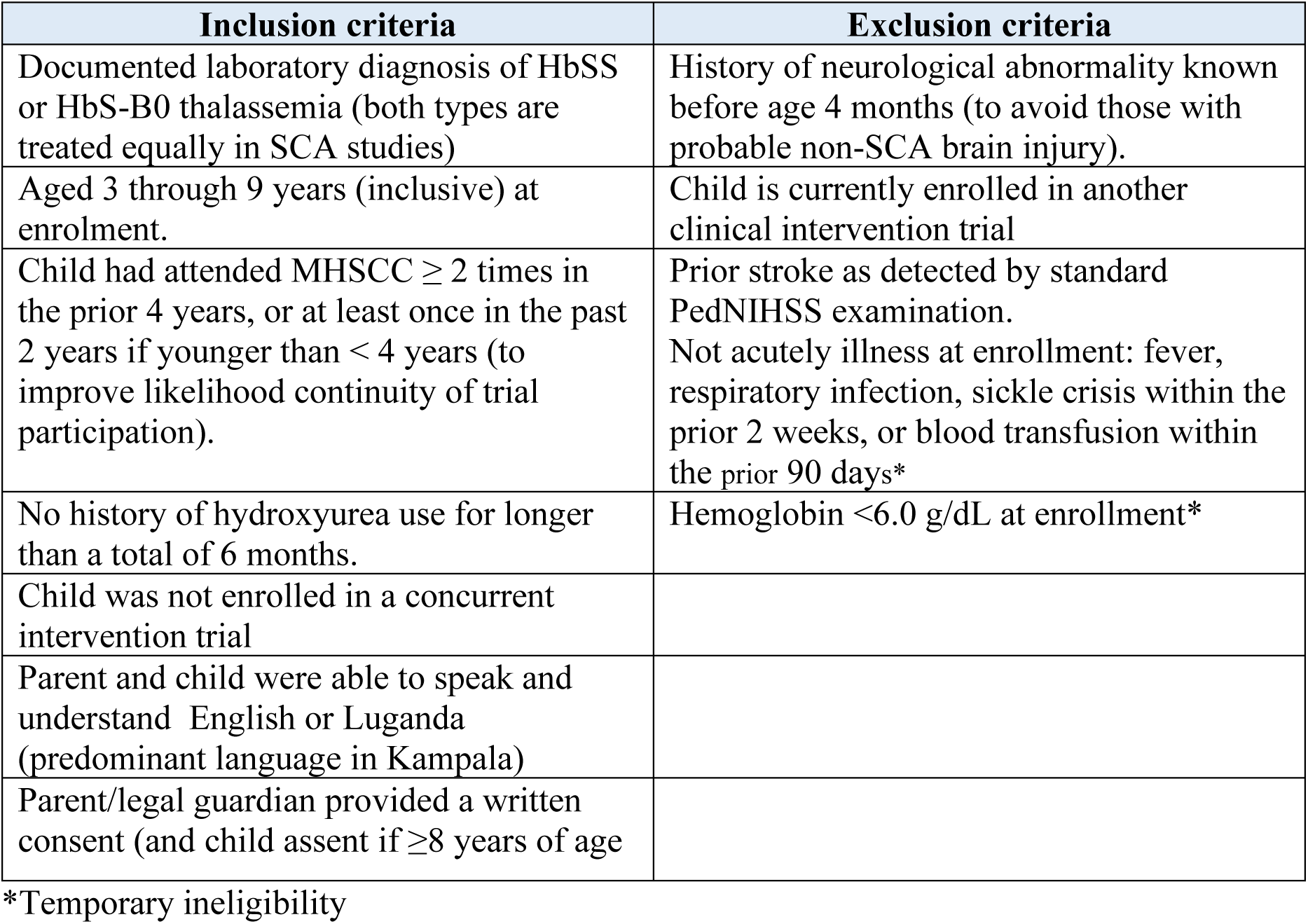
Participant eligibility criteria for participation in the BRAINSAFE II trial.

| **Inclusion criteria** | **Exclusion criteria** |
| --- | --- |
| Documented laboratory diagnosis of HbSS or HbS-B0 thalassemia (both types are treated equally in SCA studies) | History of neurological abnormality known before age 4 months (to avoid those with probable non-SCA brain injury). |
| Aged 3 through 9 years (inclusive) at enrolment. | Child is currently enrolled in another clinical intervention trial |
| Child had attended MHSCC $\geq 2$ times in the prior 4 years, or at least once in the past 2 years if younger than $< 4$ years (to improve likelihood continuity of trial participation). | Prior stroke as detected by standard PedNIHSS examination.<br>Not acutely illness at enrollment: fever, respiratory infection, sickle crisis within the prior 2 weeks, or blood transfusion within the prior 90 days* |
| No history of hydroxyurea use for longer than a total of 6 months. | Hemoglobin $< 6.0$ g/dL at enrollment* |
| Child was not enrolled in a concurrent intervention trial |  |
| Parent and child were able to speak and understand English or Luganda (predominant language in Kampala) |  |
| Parent/legal guardian provided a written consent (and child assent if $\geq 8$ years of age) | |

\*Temporary ineligibility

**Table A.2.** Primary outcomes, by trial schedule.

| **Assessments** | **Schedule** |
| --- | --- |
| Change in TCD velocity (in cm/sec);<br>Change is number in higher risk categories:<br>conditional, abnormal | At study months 0, 18, 36 |
| Neurocognitive testing by category (normal,<br>abnormal) and by mean z-scores*:<br>stable or changed (better, worse) | At study months 0, 18, 36 |
| New clinically detected stroke (yes, no) | Incident or detected at study months 18, 36 |

\*Z-scores are adjusted for age

**Table A.3.**
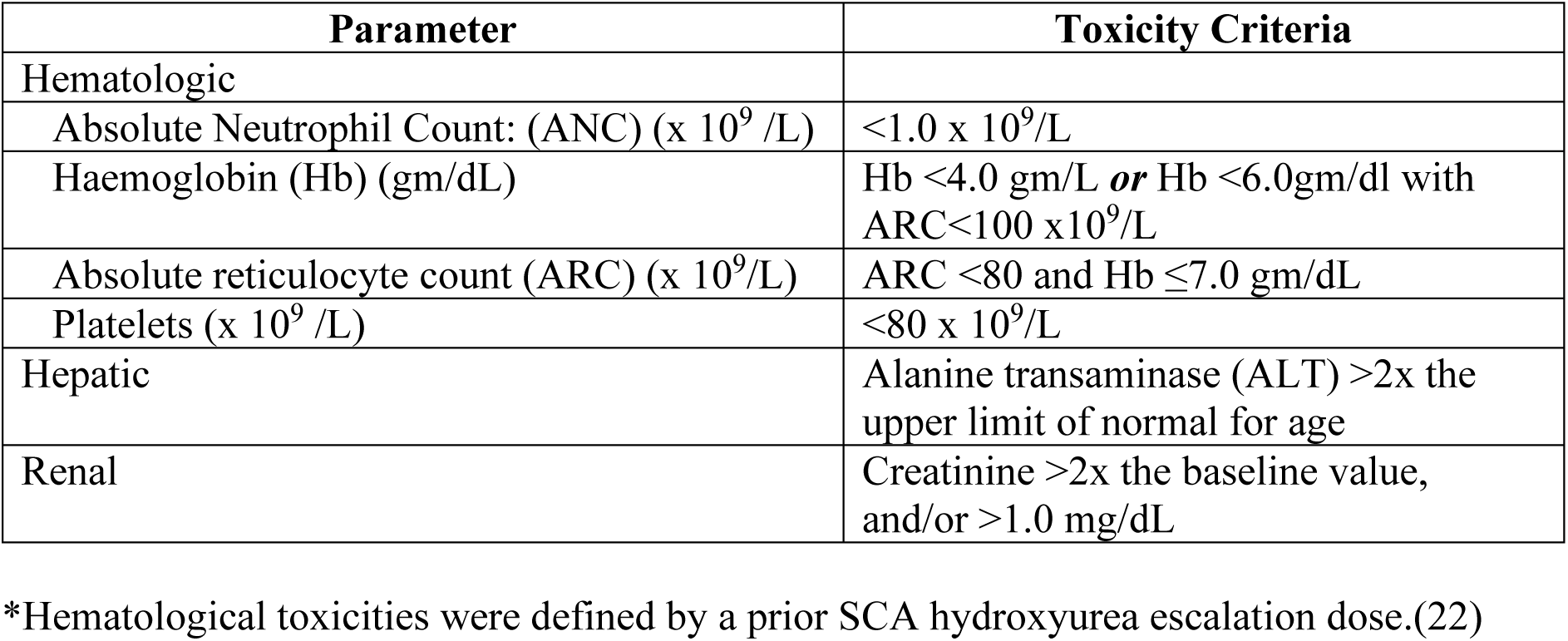
Laboratory parameters for hydroxyurea toxicity*.

| Parameter | Toxicity Criteria |
| --- | --- |
| Hematologic |  |
| Absolute Neutrophil Count: (ANC) ($\times 10^9$ /L) | $<1.0 \times 10^9$/L |
| Haemoglobin (Hb) (gm/dL) | Hb $<4.0$ gm/L *or* Hb $<6.0$gm/dl with ARC$<100 \times 10^9$/L |
| Absolute reticulocyte count (ARC) ($\times 10^9$/L) | ARC $<80$ and Hb $\leq 7.0$ gm/dL |
| Platelets ($\times 10^9$ /L) | $<80 \times 10^9$/L |
| Hepatic | Alanine transaminase (ALT) $>2$x the upper limit of normal for age |
| Renal | Creatinine $>2$x the baseline value, and/or $>1.0$ mg/dL |

\*Hematological toxicities were defined by a prior SCA hydroxyurea escalation dose.(22)

**Table A.4.** Study schedule of enrolment, interventions, and assessments.

|  | STUDY PERIOD |  |  |  |  |  |  |  |  |  |  |  |  |  |  |  |
| --- | --- | --- | --- | --- | --- | --- | --- | --- | --- | --- | --- | --- | --- | --- | --- | --- |
|  | Enrolment | Post-allocation (months) |  |  |  |  |  |  |  |  |  |  |  |  |  |  |
| TIME POINT | 0 | 1 | 2 | 4 | 6 | 8 | 10 | 12 | 15 | 18 | 21 | 24 | 27 | 30 | 33 | 36 |
| ENROLMENT: |  |  |  |  |  |  |  |  |  |  |  |  |  |  |  |  |
| Eligibility screen | X |  |  |  |  |  |  |  |  |  |  |  |  |  |  |  |
| Informed consent | X |  |  |  |  |  |  |  |  |  |  |  |  |  |  |  |
| INTERVENTION: |  |  |  |  |  |  |  |  |  |  |  |  |  |  |  |  |
| *Hydroxyurea (HU)* |  |  |  |  |  |  |  |  |  |  |  |  |  |  |  |  |
| ASSESSMENTS: |  |  |  |  |  |  |  |  |  |  |  |  |  |  |  |  |
| Clinical assessments | X | X | X | X | X | X | X | X | X | X | X | X | X | X | X | X |
| Haemoglobin Electrophoresis | X |  |  |  |  |  |  |  |  | X |  |  |  |  |  | X |
| Neurocognitive | X |  |  |  |  |  |  |  |  | X |  |  |  |  |  | X |
| PedNIHSS | X |  |  |  |  |  |  |  |  | X |  |  |  |  |  | X |
| Transcranial Doppler | X |  |  |  |  |  |  |  |  | X |  |  |  |  |  | X |
| MRI-MRA imaging (30% subset) | X |  |  |  |  |  |  |  |  |  |  |  |  |  |  | X |
| Laboratory assessments. | X | X | X | X | X | X | X | X | X | X | X | X | X | X | X | X |
| RFTs & LFTs | X |  |  |  |  |  |  | X |  |  |  | X |  |  |  | X |
| Sample storage (Plasma, RBC, CRP) | X |  |  |  |  |  |  |  |  | X |  |  |  |  |  | X |

**Fig. A. 1.**
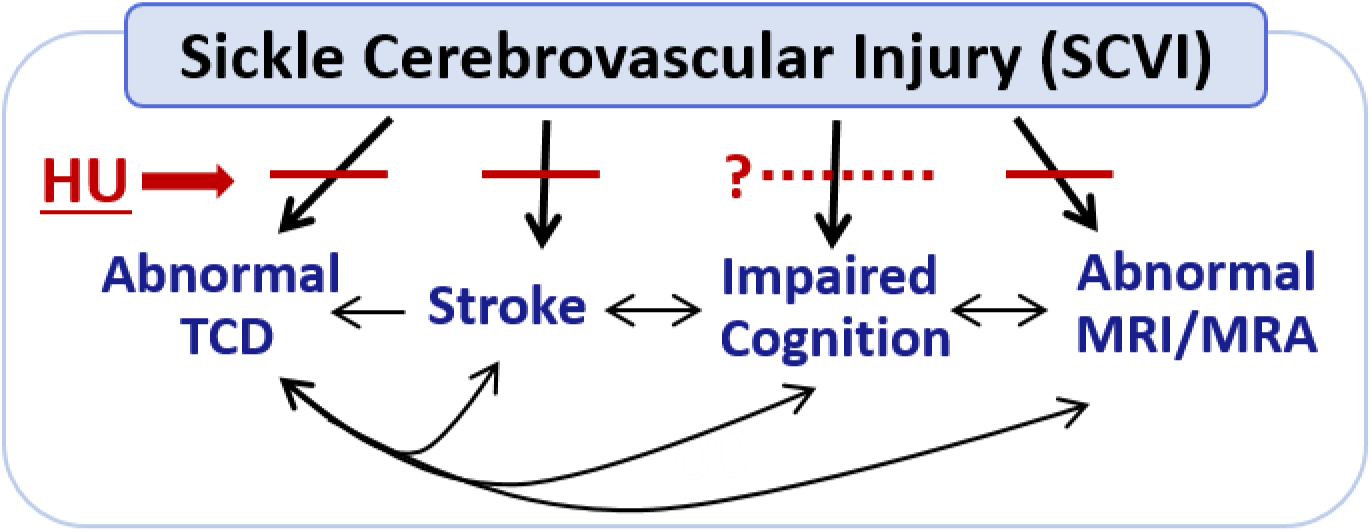
Conceptual model of hydroxyurea on sickle cerebrovascular injury (SCVI). In U.S.-based studies, hydroxyurea (HU) reduces the frequency of abnormal cranial blood flow (assessed by transcranial doppler velocity) and stroke, along with stabilization of magnetic resonance imaging, yet prospective analyses of HU effect on impaired cognition are lacking.

**Fig. A.2.**
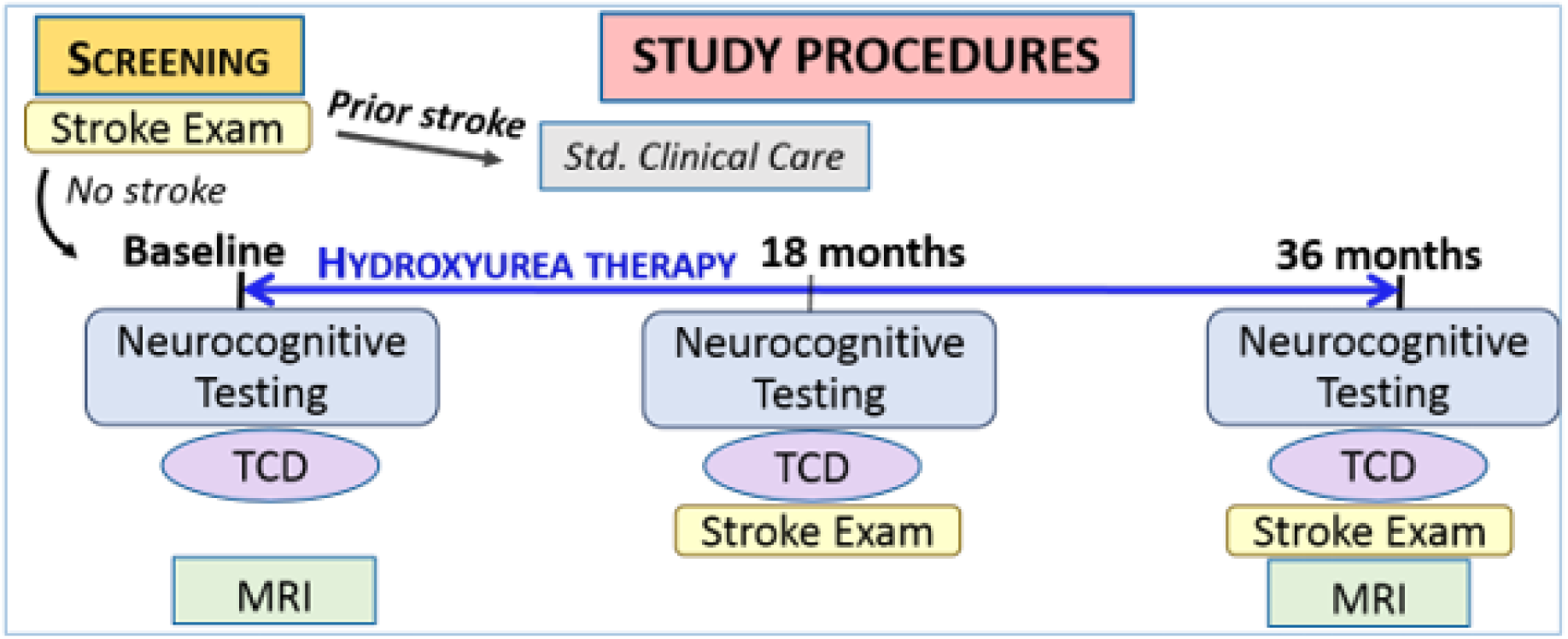
Trial scheme for treatment and scheduled endpoint assessments. Magnetic resonance imaging and angiography will be perfomed on a 30% participant sample.

**Table A.5.** BRAINSAFE II procedures for primary outcomes. All Z-scores were established by the recruited group of siblings without sickle cell anemia (SCA), adjusted for age.

| Procedure | Interpretation |  |  |
| --- | --- | --- | --- |
| Neurocognitive testing |  |  |  |
| KABC II<br>(Cognition) | Improvement | $\geq 0.5$ Z-score<sup>1</sup> | Higher composite Z-score represents better cognition/performance |
|  | Mean performance | Z-score = 0 |  |
|  | Impaired | Equal or lower than -2 Z-score |  |
| NEPSY II<br>(Executive Function) | Mean performance | Z-score = 0 | Higher composite Z-score represents better executive functioning/performance |
|  | Impaired | Equal or lower than -2 Z-score |  |
| BRIEF<br>(Executive Function) | Mean performance | Z-score = 0 | Lower composite Z-score represents fewer behavioral challenges/better executive functioning |
|  | Impaired | Equal or lower than -2 Z-score |  |
| TOVA<br>(Attention) | Mean performance | Z-score = 0 | Higher composite Z-score represents better attention |
|  | Impaired | Equal or lower than -2 Z-score |  |
| Transcranial Doppler (TCD) |  |  |  |
| Normal | < 170 cm/second |  | Categories used are internationally accepted standards for children with SCA ages 2-16 years (ref. 6) |
| Conditional | 170-199 cm/second |  |  |
| Abnormal | $\geq 200$ cm/second | | |
| Inadequate<sup>2</sup> | - |  |  |
| Pediatric NIH stroke scale (PEDNHISS) |  |  |  |
| Normal | Score = 0 |  |  |
| Stroke | Score $\geq 1$<sup>3</sup> | | |

<sup>1</sup>Approximately equivalent to a gain of 8 IQ points
<sup>2</sup>Unable to determine velocity due to poor temporal ultrasound window
<sup>3</sup>Not scored as a stroke if the child is too young to know the answer (i.e. their age or month of the year)

## Notes

Supported by award R01HD096559 (Idro, Green) from the Eunice Kennedy Shriver National Institute of Child Health and Human Development (NICHD) and the Fogarty International Center, both at the National Institutes of Health (NIH). Study drug was donated by AddMedica/Theravia, France.

### Competing Interest Statement

The authors have declared no competing interest.

### Clinical Trial

NCT04750707

### Author Declarations

Makerere University,School of medicine ethics and research committe(SOMREC).

